# Pontine Infarction: A Prospective Cohort Study on Its Clinical Features, Prognosis, and Association with Branch Atheromatous Disease

**DOI:** 10.1101/2024.01.08.24301027

**Authors:** Ryota Motoie, Kotaro Ono, Hiroshi Oketani, Yosuke Kawano, Shintaro Nagaoka, Kazushi Maeda, Yoshio Suyama, Hidefuku Gi, Yukihide Kanemoto

## Abstract

**Introduction:** This study aimed to analyse disease characteristics and prognostic factors among a cohort of 407 patients with pontine infarction, focusing on the effect of branch atheromatous disease (BAD).

**Patients and Methods:** A retrospective analysis of patients diagnosed with brainstem stroke at Baba Memorial Hospital from 2012 to 2022 was conducted. The study included patients with pontine stroke, excluding those with missing data, chronic or multiple cerebral infarctions, non-brainstem stroke, or without timely MRI. Patient analysis involved age, sex, Japan Coma Scale (JCS) score, medical history, symptoms at admission, and MRI findings. Prognostic groups were classified based on the modified Rankin Scale (mRS) and Functional Independence Measure (FIM) scores at admission and discharge.

**Results:** Of the 407 patients, 66.1% belonged to the good prognosis group (mRS 0–2) and 33.9% to the poor prognosis group (mRS 35). Younger patients tended to have a better prognosis. JCS scores correlated with prognosis severity. Significant differences in dysarthria, paralysis, and admission FIM scores were observed between patients with and without BAD. Logistic regression analysis identified the FIM score at admission as an independent predictor of prognosis.

**Discussion:** BAD was not directly related to prognosis. The absence of differences in discharge FIM scores and similar prognoses to other stroke types suggested effective rehabilitation. However, due to the lack of pre-stroke FIM measurements and specific treatment details, further research is needed.

**Conclusion:** Age and JCS scores were significant prognostic factors, with BAD not directly affecting prognosis. There is need for investigation into treatment methods and detailed prognostic factors.

## Background

Despite the availability of studies on brainstem infarction, comprehensive studies specifically focusing on pontine infarction, especially single-facility continuous case series, remain scarce. While pontine infarction generally has a relatively favourable prognosis, certain cases demonstrate increased risk and poor outcomes.^1,2^

Pontine infarction frequently involves branch atheromatous disease (BAD), a specific type of small vessel disease; however, research in this area remains inadequate. BAD primarily occurs in the basal ganglia and pons, indicating the involvement of the lenticulostriate artery (LSA) and paramedian pontine artery (PPA).^3,4^ An important event influencing prognosis is the early neurological deterioration (END), which is a characteristic feature of BAD.^5^ Hypertension, diabetes, dyslipidaemia, and smoking have been identified as risk factors for END in BAD within the LSA region; however, the individual contribution of each factor remains unclear.^6, 7^ Despite accumulating studies on the characteristics and treatment of BAD in the LSA-related basal ganglia region, research on pontine infarction involving PPA has been limited.^8, 9^

Herein, we report the results of a prospective cohort study, focusing on the characteristics, prognosis, and prognosis-determining rehabilitation aspects of pontine infarction inclusive of BAD.

## Patients and Methods

### patients

We identified 751 patients diagnosed with brainstem stroke, including midbrain, pons, and medulla oblongata infarctions, from the electronic medical records of the Baba Memorial Hospital(Sakai-shi, Japan) from 2012 to 2022. We excluded cases with missing data (26 cases), chronic brainstem infarctions (26 cases), multiple cerebral infarctions, including basilar artery occlusions (62 cases), non-brainstem stroke (92 cases), cases without magnetic resonance imaging (MRI) within 3 d of hospitalisation (38 cases), and asymptomatic cases (15 cases). Finally, a focused analysis was conducted on 407 pontine stroke cases, excluding 84 non-pontine brainstem stroke cases (Figure 1). All medical records were stored in the electronic database of the Baba Memorial Hospital. The institutional clinical medical ethics committee of Baba Memorial Hospital approved this study (2024-01). All patients or their relatives provided written informed consent before treatment.

**Figure 1.**
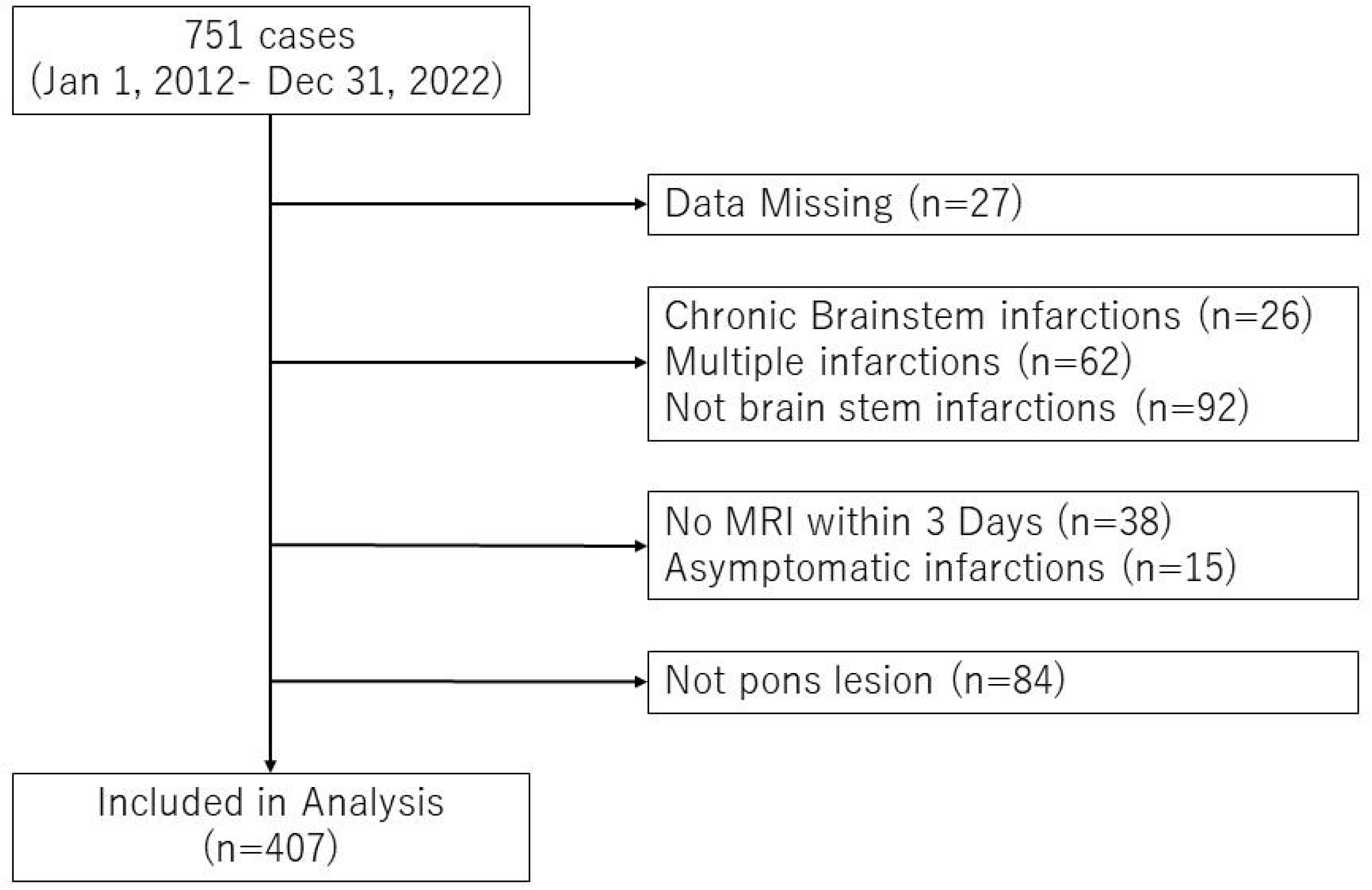
Flow diagram of the study selection process.

### Study Design and Population

The collected clinical information of patients included age, sex, initial Japan Coma Scale (JCS) score, medical history (hypertension, diabetes mellitus, dyslipidaemia, and stroke), history of antithrombotic drug use, symptoms at admission (ataxia, dysarthria, and hemiparesis), smoking history, daily drinking habits, and magnetic resonance imaging (MRI) findings indicating BAD at or around the time of admission. The discharge modified Rankin Scale (mRS) and functional independence measure (FIM) scores at admission and discharge were also obtained.

BAD was defined as high signal intensity in the basal part of the pons on diffusion-weighted imaging.^10^ Prognostic groups were classified based on discharge mRS scores, with a mRS score of 0–2 indicating favourable prognosis and a score of 3–6 indicating poor prognosis.^11^

FIM scores were assessed by clinicians at admission and discharge. The FIM scoring system was developed in 1983 by the American Congress of Rehabilitation Medicine and the American Academy of Physical Medicine and Rehabilitation^12^ and comprises 13 items assessing physical function and five items evaluating cognitive function; each item is scored from 1 to 7. This scoring system facilitates the precise assessment of the daily activities of a patient, making it a frequently used tool for determining the effectiveness of rehabilitation and predicting outcomes.^13,14^

In this study, patients with brainstem infarction were stratified into groups with favourable and poor prognoses. Univariate analysis was conducted to delineate the background characteristics of each group. Subsequently, based on previous studies, logistic regression analysis was utilised to assess the extent to which various prognostic factors influence the likelihood of patient outcomes.^15–19^

In addition, this study aimed to determine whether pontine infarction with BAD leads to worse prognosis, similar to cases with BAD in the basal ganglia, by comparing patients with and without BAD. To compare patient backgrounds, variables such as age, sex, medical history (hypertension, diabetes, dyslipidaemia, and stroke), lifestyle habits (smoking and regular alcohol use), symptoms at admission (paralysis, ataxia, and dysarthria), length of hospital stay, and FIM scores at admission and discharge were analysed. Given the lack of clarity on risk factors for BAD, multivariate analysis was conducted to identify independent risk factors, based on previous studies.^5,20^

Most patient data are expressed as categorical variables, except for age and FIM scores, which are expressed as continuous variables. All groups were tested using Fisher’s exact test for categorical variables or Mann-Whitney U test for continuous variables. All statistical analyses were performed using R (version 4.3.2; R Foundation for Statistical Computing, Vienna, Austria). A P-value of <0.05 was considered statistically significant. This study was approved by the Ethics Committee of Baba Memorial Hospital and was reported following the Strengthening of Reporting of Observational Studies in Epidemiology statement.

## Results

Comparison between Groups with Good and Poor Prognosis

In this analysis of 407 patients with brainstem stroke, 269 (66.1 %) were classified in the good prognosis group (mRS 0–2) and 138 (33.9 %) in the poor prognosis group (mRS 3–5). Notably, we observed that patients in the good prognosis group were younger, with a median age of 72 years (interquartile range [IQR], 65–79 years), than those in the poor prognosis group, with a median age of 79 years (IQR, 70–85 years) (p < 0.001).

When examining prognosis in relation to JCS scores, we detected that 24.6 % of patients with a JCS of 0 were in the good prognosis category, whereas 53.5 % were in the poor prognosis category. We observed a trend in which an increase in JCS score corresponded to a decrease in the rate of good prognosis and an increase in the rate of poor prognosis. This was especially pronounced in patients with high JCS scores, who exhibited a markedly lower rate of good prognosis and a significantly higher rate of poor prognosis (p < 0.001).

In terms of medical history, we identified that the prevalence of hypertension, dyslipidaemia, and diabetes was not significantly different between the two groups. However, we noticed that a prior history of stroke was more prevalent in the poor prognosis group (20.4 % vs 34.1 %, p = 0.004). With respect to symptoms at admission, we did not observe any significant differences in the occurrence of dysarthria and ataxia between the groups. Nevertheless, the incidence of paralysis was notably lower in the good prognosis group (53.9 % vs 65.9 %, p = 0.02) (Table 1).

**Table 1.** Comparative Analysis of Favorable versus Unfavorable Outcomes in in Pontine Infarction.

|  | All patients | Good prognosis<br>(mRS 0-2) | Poor prognosis<br>(mRS 3-5) | *p* Value |
| --- | --- | --- | --- | --- |
| Number (%) | 407 | 269 (66.1) | 138 (33.9) |  |
| Sex (%), male | 278 | 87 (63.0) | 191 (71.0) | 0.115 |
| Age, median (IQR) | 76 (54-93) | 72 (65-79] | 79 (70-85) | <0.001 |
| BAD | 119 | 70 (26.0) | 49 (35.5) | 0.660 |
| JCS (%) |  |  |  |  |
| 0 |  | 34 (24.6) | 144 (53.5) | <0.001 |
| 1 |  | 99 (36.8) | 42 (30.4) |  |
| 2 |  | 17 ( 6.3) | 34 (24.6) |  |
| 3 |  | 8 ( 3.0) | 19 (13.8) |  |
| 10 |  | 1 ( 0.4) | 5 ( 3.6) |  |
| 20 |  | 0 ( 0.0) | 1 ( 0.7) |  |
| 100 |  | 0 ( 0.0) | 1 ( 0.7) |  |
| 200 |  | 0 ( 0.0) | 2 ( 1.4) |  |
| Hypertension (%) | 266 | 178 (66.2) | 88 (63.8) | 0.914 |
| Diabete mellitus (%) | 161 | 108 (40.1) | 53 (38.4) | 0.749 |
| Dyslipidemia (%) | 76 | 56 (20.8) | 20 (14.5) | 0.140 |
| pre-Stroke (%) | 102 | 55 (20.4) | 47 (34.1) | 0.004 |
| Smoking history (%) | 192 | 62 (44.9) | 130 (48.3) | 0.531 |
| Daily alcohol use (%) | 136 | 100 (37.2) | 36 (26.1) | 0.027 |
| Antithrombotic drugs (%) | 113 | 68 (25.3) | 45 (32.6) | 0.129 |
| Paralysis (%) | 236 | 145 (53.9) | 91 (65.9) | 0.020 |
| Ataxia (%) | 25 | 18 ( 6.7) | 7 ( 5.1) | 0.664 |
| Dysarthria (%) | 262 | 175 (65.1) | 87 (63.0) | 0.83 |

BAD: Branched atheromatous disease; JCS: Japan Coma Scale; IQR: Interquartile range.

In logistic regression analysis, we considered various factors as independent variables: age, sex, presence of BAD, hypertension, diabetes, dyslipidaemia, smoking history, habitual alcohol consumption, and FIM score at admission. Our analysis revealed that the FIM score at admission was the sole independent predictive factor (admission FIM odds ratio [OR] = 1.060, 95 % confidence interval [CI] = 1.050–1.070) (Table 2). Utilising this model to determine the optimal cutoff point (cutoff = 0.568), we identified that the admission FIM threshold was 60.39. This threshold demonstrated a sensitivity of 93.0 % and specificity of 76.0 % in predicting prognosis (Figure 2).

**Table 2.** Multivariate Analysis of Variables in Patients with Good prognosis versus Poor prognosis.

|  | **OR** | **SE** | **95% CI** | **p-value** |
| --- | --- | --- | --- | --- |
| Age | 0.979 | 0.014 | 0.953-1.01 | 0.134 |
| Sex | 1.330 | 0.319 | 0.71-2.48 | 0.376 |
| BAD | 0.800 | 0.286 | 0.457-1.4 | 0.436 |
| Hypertension | 0.937 | 0.298 | 0.523-1.68 | 0.827 |
| Diabetes mellitus | 0.960 | 0.286 | 0.548-1.68 | 0.888 |
| Dyslipidemia | 1.310 | 0.368 | 0.638-2.7 | 0.460 |
| Smoking history | 0.657 | 0.317 | 0.353-1.22 | 0.185 |
| Daily alcohol use | 0.920 | 0.308 | 0.503-1.68 | 0.787 |
| Admission FIM | 1.060 | 0.007 | 1.05-1.08 | <0.001 |

OR: Odds ratio; SE: Standard error; CI: Confidence limit; BAD: Branched atheromatous disease; JCS: Japan Coma Scale; FIM: Functional independence measure.

**Fig. 2.**
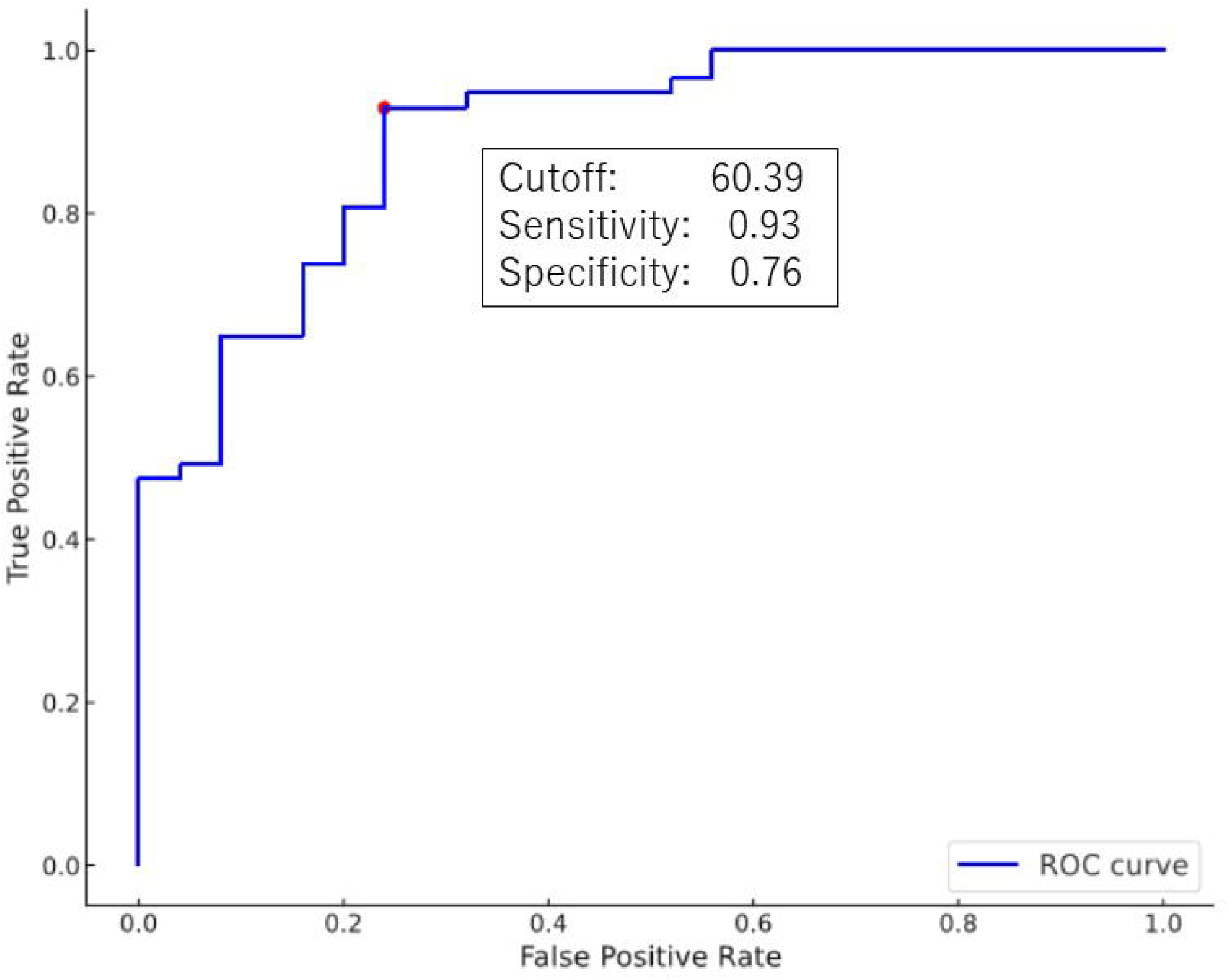

In BAD analysis, we identified that out of the total cohort, 119 (29.2 %) patients had BAD, whereas 288 (70.8 %) did not exhibit BAD. Our analysis revealed no significant differences in age and sex between the BAD and non-BAD groups. Notably, we observed that the BAD group presented a higher incidence of paralysis and dysarthria compared with those in the non-BAD group (paralysis: 70.6 % vs 52.8 %, p = 0.001; dysarthria: 76.5 % vs 59.4 %, p = 0.001). Additionally, we determined that the FIM score at admission was significantly lower in the BAD than in the non-BAD group (p < 0.001). However, we did not observe any differences in the discharge FIM scores or duration of hospital stay between the two groups. The results of mRS comparison between BAD and non-BAD groups showed no significant differences (Figure 3).

**Fig. 3.**
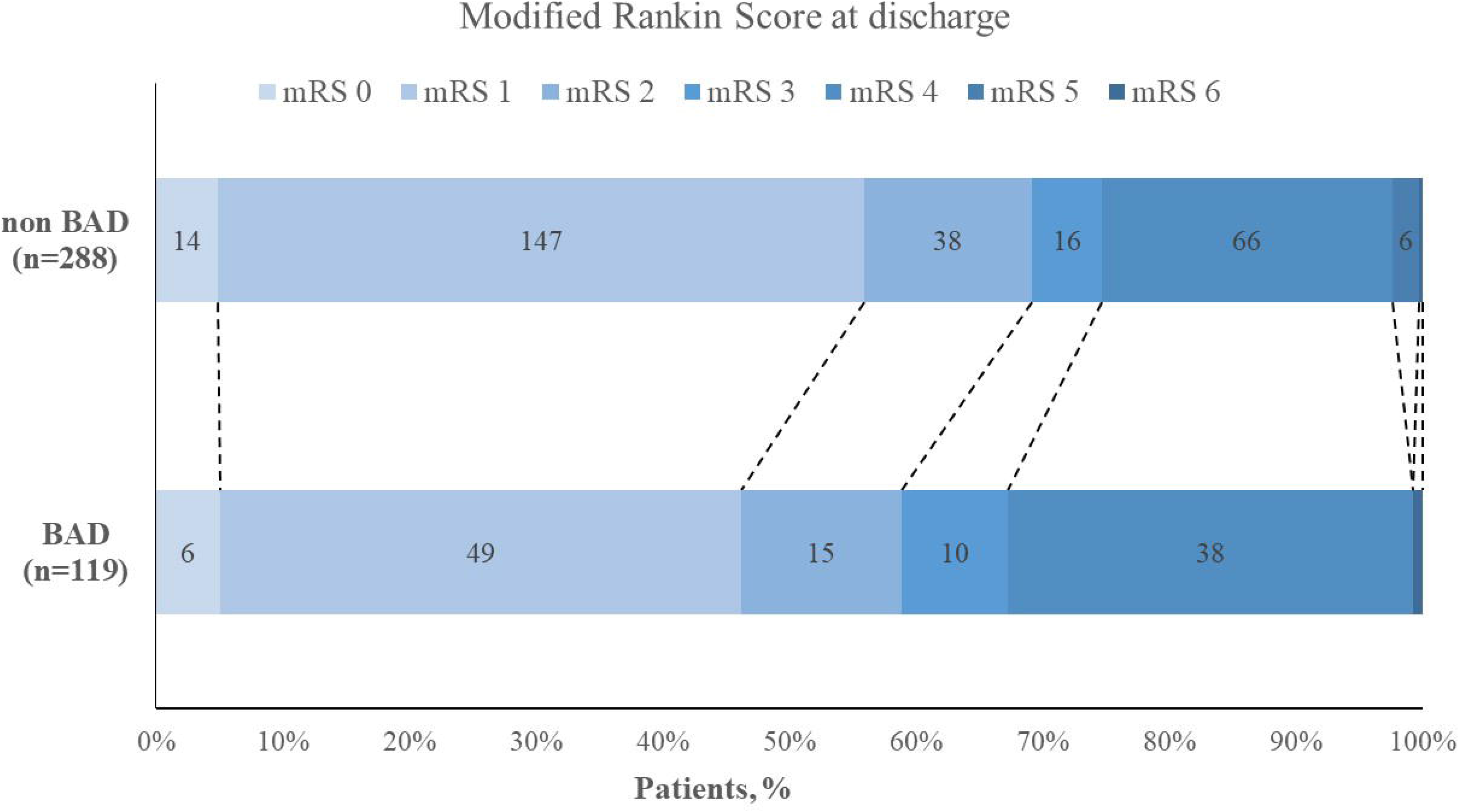

In the multivariate analysis, we evaluated variables such as age, underlying diseases, and presenting symptoms as independent factors for the occurrence of BAD. However, we did not definitively find a direct association of these factors with the development of BAD (Table 3).

**Table 3.**
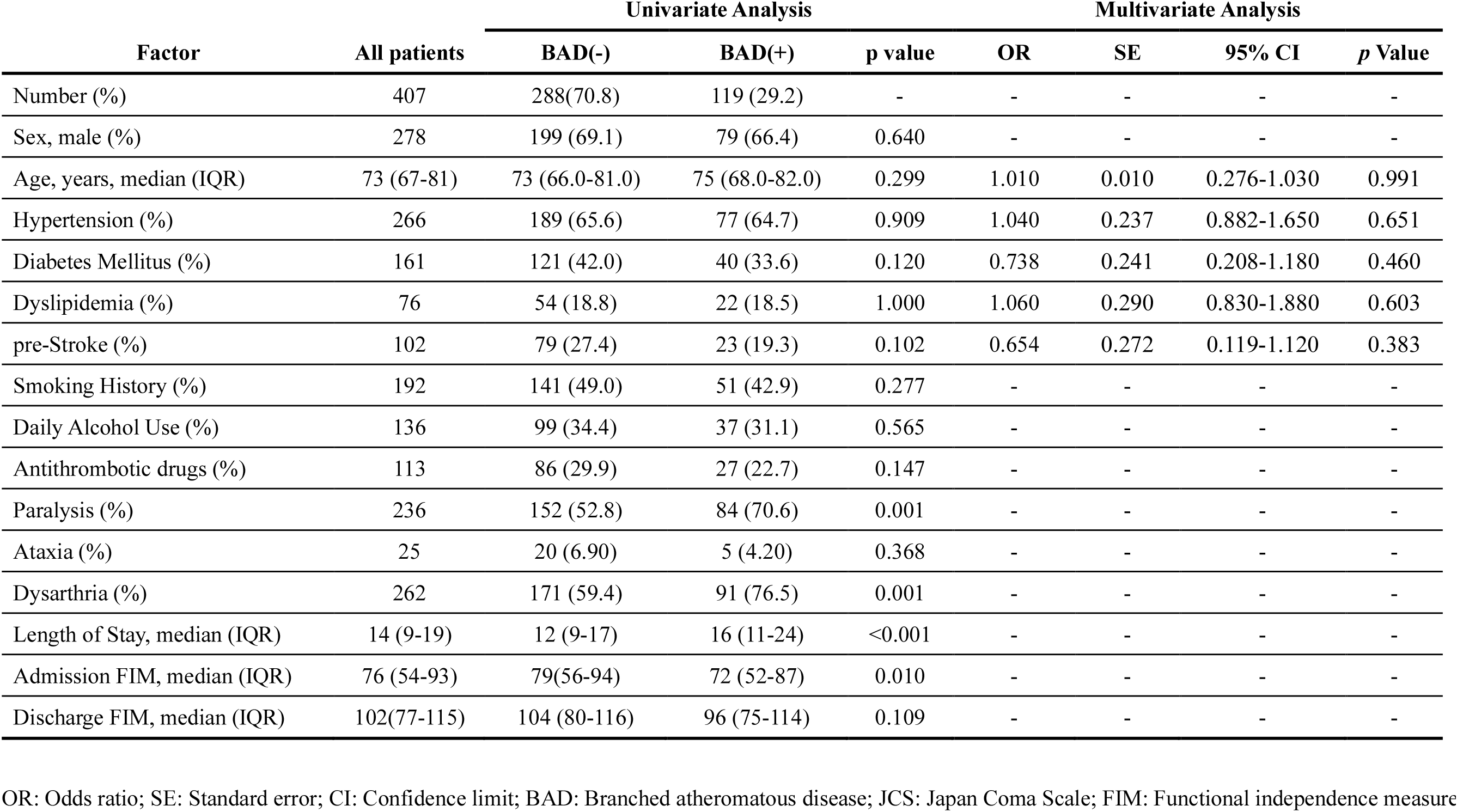
Multivariate Analyses of Variables in Patients with Brainstem Auditory Dysfunction (BAD) Versus Non-BAD.

## Discussion

The cohort of 407 patients in this study was divided into different prognosis groups, with 66.1 % classified into the good prognosis category (mRS 0–2) and 33.9 % into the poor prognosis category (mRS 3–5), as shown in Table 1. A statistically significant younger median age was observed in the good prognosis group (72 years) than that in the poor prognosis group (79 years) (p < 0.001). This suggested a tendency for younger patients to have a better prognosis. Reflecting upon the findings of a study from 2016, which indicated that approximately 63 % of patients with stroke over the age of 70 were discharged home, our data implied that the prognosis for patients with pontine infarction may be similar to, or potentially better than that for patients with other types of strokes.^21^

Analysis of the JCS scores revealed a trend in which an increase in severity was associated with a higher likelihood of poor prognosis, indicating that the JCS score is a reliable indicator of stroke severity. Regarding medical history, we observed that the incidences of hypertension, dyslipidaemia, and diabetes were not significantly different between groups. However, it is noteworthy that a history of stroke was more commonly observed in the poor prognosis group (p = 0.004). Among admission symptoms, hemiplegia emerged as a significant factor influencing prognosis (p = 0.02), highlighting the need for further investigation into the relationship between stroke history and hemiplegia in future studies.

Several factors were considered in the logistic regression analysis, with the FIM score at admission standing out as an independent predictor of prognosis (OR = 1.060, 95 % CI = 1.050– 1.070). Nevertheless, caution is warranted in interpreting these results due to the lack of pre-stroke FIM score measurements. This suggested a probable trend in which patients with more severe conditions are likely to experience poorer prognoses; though further validation is necessary. The study model calculated a cutoff point for the admission FIM score of 60.39, exhibiting a sensitivity of 93.0 % and a specificity of 76.0 %. This finding implied that an admission FIM score above 61 could serve as a useful independent predictor of prognosis; however, additional investigations are required to verify its potential in clinical application.

The general prognosis of BAD is typically poor, as established in prior studies.^9,12^ Consequently, this study posited that BAD might contribute to a poor prognosis in pontine infarction cases. Contrary to this hypothesis, the findings of this study revealed no correlation between BAD and prognosis. For a more nuanced understanding of BAD, patients were categorised into two groups based on the presence or absence of BAD. Subsequent univariate analysis yielded significant differences in the prevalence of dysarthria and paralysis, as well as in the FIM scores at admission, between patients with and without BAD (Table 3). However, no significant differences were observed in the FIM scores at discharge between the two groups. This indicated that while dysarthria and paralysis in pontine infarction with BAD may contribute to lower FIM scores at admission, these disparities tend to diminish by discharge, likely due to effective rehabilitation. Given the scarcity of specific literature on pontine infarction, direct comparison with existing studies is challenging.

Nonetheless, this investigation, focusing on BAD in the pontine region, suggested that the presence of BAD does not influence prognosis, aligning with the results of previous studies that based prognosis on MRI findings.^22^ This outcome could be hypothesised to result from successful BAD treatment in the pontine region; however, as the treatment aspect was not specifically examined in this study, this explanation remains speculative. Further research is warranted to delve into these aspects.

## Conclusion

This study, which focused on prognostic factors in a cohort of 407 patients with stroke, identified age as a significant determinant. Additionally, analysis of the JCS scores revealed a correlation between higher severity and poorer prognosis. Notably, the presence or absence of BAD did not directly impact prognosis, suggesting the potential effectiveness of rehabilitation. However, the specifics of the treatment methods used remain undefined, underscoring the necessity for further investigation in this area.

## Limitations

The treatment of pontine infarction was not specifically investigated; therefore, potential variations in treatment modalities and their outcomes remain unexplored. Additionally, the impact of END on prognosis has not been established due to a lack of research in this area. Consequently, further research is essential to elucidate the characteristics and prognosis of pontine infarction, particularly in relation to BAD.

## Data Availability

All data produced in the present study are available upon reasonable request to the authors

## Declarations

### Informed consent and Ethical approval

The institutional clinical medical ethics committee of Baba Memorial Hospital approved this study (2024-01). All patients or their relatives provided written informed consent before treatment.

### Guarantor

**RM**

### Contributorship

RM wrote the first draft of the manuscript. All authors reviewed and edited the manuscript and approved the final version of the manuscript.

## Acknowledgments

We express our profound gratitude to the SCU staff and Medical Information Management Department of the Baba Memorial Hospital.

